# Diabetic retinopathy environment-wide association study (EWAS) in NHANES 2005-8

**DOI:** 10.1101/2020.09.20.20198218

**Authors:** Kevin Blighe, Sarega Gurudas, Ying Lee, Sobha Sivaprasad

**Author notes:** corresponding author: Professor Sobha Sivaprasad.

## Abstract

**Background:** Several circulating biomarkers are reported to be associated with diabetic retinopathy (DR). However, their relative contributions to DR compared to known risk factors, such as hyperglycemia, hypertension, and hyperlipidemia, remain unclear. In this data driven study, we used novel models to evaluate the associations of over 400 laboratory parameters with DR.

**Methods:** We performed an environment-wide association study (EWAS) of laboratory parameters available in National Health and Nutrition Examination Survey (NHANES) 2007-8 in individuals with diabetes with DR as the outcome (test set). We employed independent variable (‘feature’) selection approaches, including parallelized univariate regression modeling, Principal Component Analysis (PCA), penalized regression, and RandomForest™. These models were replicated in NHANES 2005-6 (replication set).

**Findings:** The test and replication set consisted of 1025 and 637 individuals with available DR status and laboratory data respectively. Glycohemoglobin (HbA1c) was the strongest risk factor for DR. Our PCA-based approach produced a model that incorporated 18 principal components (PCs) that had AUC 0.796 (95% CI 0.761-0.832), while penalized regression identified a 9-feature model with 78.51% accuracy and AUC 0.74 (95% CI 0.72-0.77). RandomForest™ identified a 31-feature model with 78.4% accuracy and AUC 0.71 (95% CI 0.65-0.77). On grouping the selected variables in our RandomForest™, hyperglycemia alone achieved AUC 0.72 (95% CI 0.68-0.76). The AUC increased to 0.84 (95% CI 0.78-0.9) when the model also included hypertension, hypercholesterolemia, hematocrit, renal and liver function tests.

**Interpretation:** All models showed that the contributions of established risk factors of DR especially hyperglycemia outweigh other laboratory parameters available in NHANES.

**RESEARCH IN CONTEXT:** What is already known about this subject?

▪ There are >500 publications that report associations of candidate circulating biomarkers with diabetic retinopathy (DR).
▪ Although hyperglycemia, hypertension, and hyperlipidemia are established risk factors, they do not always explain the variance of this complication in people with diabetes; DR also shares risk factors with other diabetes complications including markers of renal and cardiovascular disease.
▪ ‘Holistic’ studies that quantify risk across all of these parameters combined are lacking.

What is the key question?

▪ It is unclear whether risk models for DR may be improved by adding some of these reported biomarkers - there is an unmet need to systematically evaluate as many circulating biomarkers as possible to help rank their associations with DR.

What are the new findings?

▪ We show that hyperglycemia is the strongest risk factor across all models.
▪ We stratified the rest of the highest ranked parameters into groups related to diabetes control, renal and liver function, and hematocrit changes.

How might this impact on clinical practice in the foreseeable future?

▪ The importance of focusing on parameters beyond hyperglycemia control to reduce risk of progression from diabetes to DR is emphasized.

## INTRODUCTION

Diabetes represents the most common cause of microvascular changes in the retina. The initial retinal lesions of diabetic retinopathy (DR) are microaneurysms but they can occur in eyes with and without diabetes (1-3). With increasing duration of diabetes, other lesions develop and co-exist in the retina such as retinal hemorrhages, exudates, intraretinal microvascular abnormalities and neovascularization of the retina or optic disc. Based on the presence of individual lesions or a constellation of them, DR severity level is graded from mild, moderate and severe non-proliferative diabetic retinopathy (NPDR) to proliferative diabetic retinopathy (PDR) (4, 5). Diabetic macular edema (DME) can occur in any stage of DR (5). In population-based studies, approximately a third of people with diabetes have DR (6, 7). The established systemic risk factors for DR are suboptimal control of hyperglycemia, hypertension and hyperlipidemia (8, 9). Hypertension can also cause some of these retinal lesions independent of diabetes (10).

There are several laboratory parameters that have been shown to be abnormal in people with DR such as hyperuricemia (11), low vitamin D levels (12), low thyroxine levels (13), anemia (14), oxidative stress and inflammatory markers (15). In addition, DR is also associated with markers of diabetic kidney disease including microalbuminuria and serum creatinine (16, 17) and cardiovascular disease markers such as raised C-reactive protein (CRP) (18). Most of these associations and risks of DR are reported based on analysis of candidate laboratory-based serum or urinary markers.

In addition to these risk factors, there are several other non-modifiable and modifiable risk factors that have been attributed to the development and progression of DR. Some of these include age of onset of diabetes, duration of diabetes, male sex, and ethnicity (19-21).

There is an unmet need to rank these reported retinal, systemic and laboratory risk factors to understand their relative contributions or associations with DR in people with diabetes. The National Health and Nutrition Examination Survey (NHANES - https://wwwn.cdc.gov/Nchs/Nhanes/) was initiated in the 1960s in order to examine the health and nutritional status of US citizens and has been surveying the population up to the present time. Since 1999, it has examined ∼5000 citizens per year and includes various topics, including cardiovascular disease, diabetes, environmental exposures, eye diseases, hearing loss, infectious diseases, kidney disease, nutrition, etc. The data also contains several laboratory markers including environmental toxins, allergens, pollutants.

In this study, we used an environment wide association study (EWAS) methodology (22-25) on NHANES 2007-8 to evaluate the rank order of systemic and laboratory risks of DR among individuals identified as having diabetes to evaluate their relative associations with DR. Our findings are then replicated in NHANES 2005/6. Our objective is not to only use previously reported risk factors but also provide new research avenues from this data driven agnostic modelling study.

## METHODS

### Study data preparation

We used National Health and Nutrition Examination Survey (NHANES) 2007-8 as our primary cohort and 2005-6 as a replication cohort. Both datasets were prepared in the same fashion, however, for ease of interpretation, the following methods describe 2007-8. Specifically, three main categories of data were used: examination data (*Ophthalmology - Retinal Imaging data*; OPXRET_E), demographics data (DEMO_E), and laboratory data (**Figure 1** footnote). The main outcome of interest in the examination data was *4 levels retinopathy severity, worse eye* (OPDURL4) – this variable was recoded as binary with levels: no retinopathy; retinopathy (including mild NPR, moderate/severe NPR, and proliferative). All datasets were downloaded as SAS XPORT (xpt) format and read into R (v4.0.2) via the *Hmisc* package.

**Figure 1.**
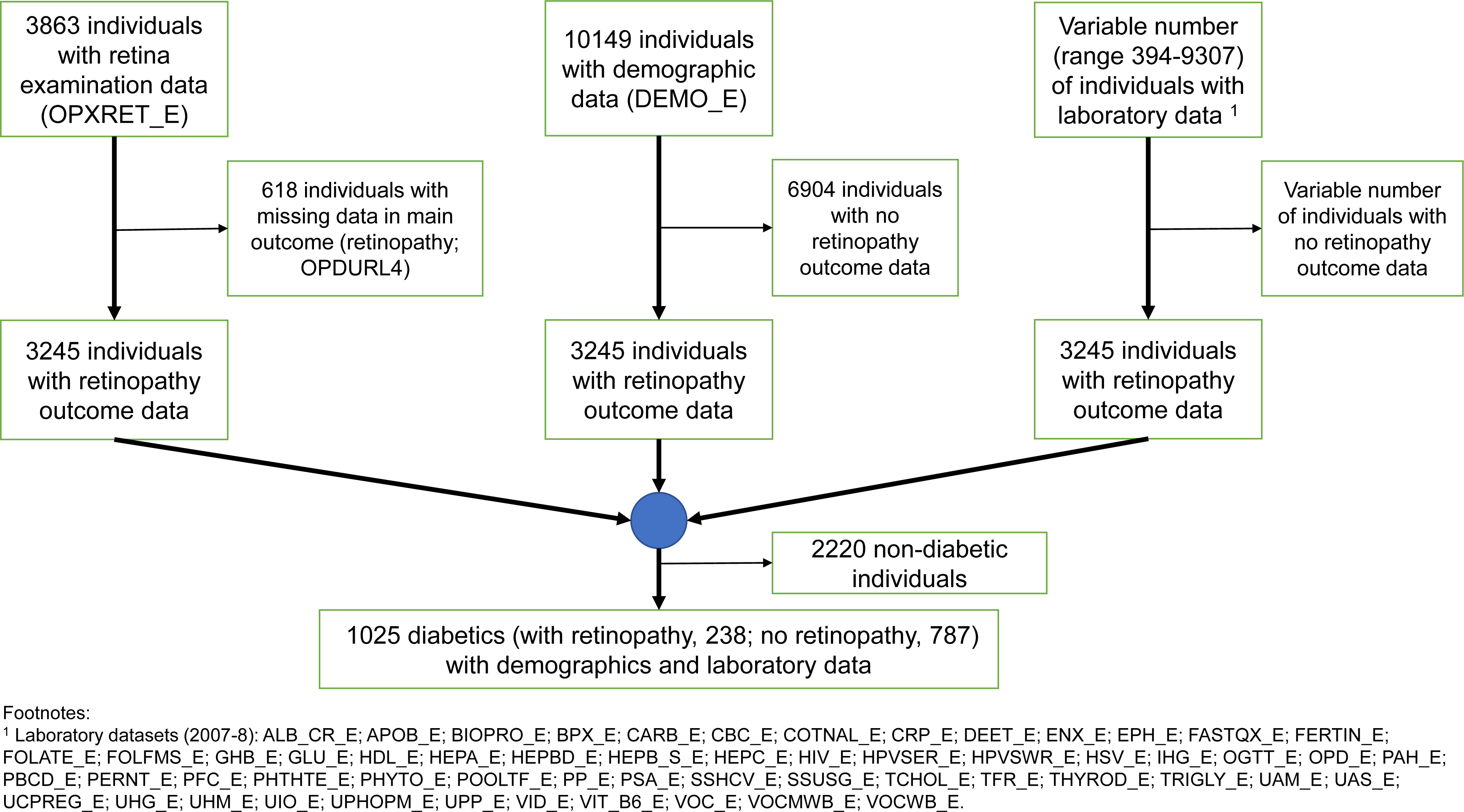
Cohort selection process for NHANES 2007-8.

Individuals with a missing value in the main outcome variable were removed before aligning the examination, demographics, and laboratory data via each individual’s respondent sequence number (SEQN). This dataset was then further filtered for only those individuals who had diabetes (**Figure 1**). Variables were removed from the data that had 0 variance (i.e. constant values) (**Supplementary Table 1**). Prior to any analysis, in addition, any variable that contained a single value occupying > 90% of total values was removed, as were variables that had > 90% missingness. Further specific filtering and encoding was then applied per dataset. [A] Examination data: variables that were different encodings of the main outcome were removed; variables that related to the status of the examination appointment were removed; OPDUHMA was removed, as it is a combination of 2 other variables that were retained (OPDUMA and OPDUHEM); variables related to glaucoma, for which there is already a single variable, were removed; variables related to the left or right eye where there was already a variable for ‘worse’ eye were removed; values encoded as missing were recoded as NA; and all other remaining variables were encoded as binary, with 0 representing the absence of the condition, and 1 representing any recorded presence (at any level) of the condition. [B] Demographic data: variables associated with interpreters and the language of the interview were removed; variables that were duplicates or different encoding of each were removed. [C] Laboratory data: categorical variables were removed and only continuous retained; duplicate variables related to the oral glucose tolerance test (OGTT_E) were removed; variables related to time since domestic activities (‘pump gas’, ‘shower’, etc.) were removed; variables that were duplicates or different encodings of each were removed; variables measured on the imperial system of weights and measures were removed if they had a corresponding variable in SI units. We focused only on continuous laboratory variables for the following reasons: 1, in NHANES, the majority of categorical variables are derived from the continuous variables; 2, our PCA-based approach can only work on continuous variables; 3, for RandomForest™, having continuous variables increases the number of splitting points in the data, and metrics of importance such as Gini are known to exhibit less bias on such data (26).

### Diabetes status

To define the diabetes status for each individual, questionnaire data (DIQ_E) was used in addition to variables already included in the laboratory data. Diabetes status was then defined as an individual satisfying any of the following: Self-reported diabetes (DIQ010); on anti-diabetes drugs (DIQ070); taking insulin (DIQ050); fasting blood sugar (FBS) ≥ 6.1 (110mg/dl) (LBDGLUSI); random blood sugar (RBS) ≥ 11.1 (200mg/dl) (LBDSGLSI); oral glucose tolerance test (OGTT) ≥ 200mg/dl (LBDGLTSI); Glycohemoglobin (HbA1c) ≥ 6.5% (LBXGH).

### Covariates

Age, ethnicity, and diabetes duration were used as covariates. Diabetes duration was calculated as age at screening minus the age at which the individual was first informed that he/she had diabetes.

### Statistical analysis

Prior to statistical analysis, continuous laboratory variables were logged (log_e_) and then transformed into z-scores to ensure that these were on the same scale. In regression analysis, the complex sampling design of the NHANES dataset was accounted for through use of survey sampling weights via the *survey* package in R / CRAN. To do this, the following value-pairs were used with the *svydesign* function: (*id*, SDMVPSU; *strata*, SDMVSTRA; *weights*, WTMEC2YR; *nest*, TRUE).

Univariate analysis was performed on all candidate predictors using a survey-weighted compute-parallelized logistic regression model via the R / Bioconductor package *RegParallel*, adjusting for age, ethnicity, and duration of diabetes separately. The Benjamini-Hochberg (27) procedure was used to control the type I error false discovery rate (FDR). A customized Manhattan plot was generated using *ggplot2*, while pairwise scatter and correlation plots were generated via a customized pairs plot. Finally, a heatmap was generated via the R / Bioconductor package *ComplexHeatmap*.

As our study is also hypothesis-generating, multivariate approaches based on principal component analysis (PCA), penalized regression, and the RandomForest™ classification algorithm were additionally used. Variables were pre-filtered and prepared as per univariate testing. Principal component analysis was performed via the R / Bioconductor package *PCAtools*. After conducting PCA, each eigenvector was then independently regressed against retinopathy outcome via binary logistic regression and those that passed p≤0.05 were used to construct a multivariable model that was further tested in ROC analysis via the *pROC* package in R.

Separately, as model complexity and multi-collinearity can arise from a large number of predictors, elastic net regularization (penalized regression with L1 and L2 penalties of the Lasso and Ridge methods) was used to reduce the number of predictor variables using *glmnet* in R / CRAN. To fit the model, 100x cross-validation was used and alpha (α) set to 0.5. The final chosen variables were those whose coefficients were not shrunk to zero – these were plot as violin plots with scatter overlays to show differences between non-DR and DR via *ggplot2*. To determine accuracy, model predictions were made on the data using the lambda (λ) one-standard-error rule using the *predict* function from the *stats* package in R.

Finally, the RandomForest™ (RF) model was fitted via the *randomForest* R / CRAN package. For this, the dataset was divided randomly into 50% training and 50% validation. Prior to model fitting, the initial model was tuned using functionality provided by the *caret* package in R / CRAN, as follows: 1), a 10x cross-validation control function was defined via *trainControl* function; 2) the best value for ‘mtry’, i.e., the ideal number of variables to randomly sample, was determined using the *train* function across a search / tuning grid ranging between 1-40 and with Kappa as the metric; and 3) using the selected value of ‘mtry’, the ideal number of trees, ‘ntrees’, was determined also via the *train* function with selection metric based on Kappa. After the initial model was fit, variables with mean decrease in accuracy ≤ 1% were excluded and the model re-fit. This was then repeated in a recursive fashion until all variables with negative mean decrease accuracy were removed from the model.

### Final risk models

Variables selected from RandomForest™ were grouped based on similarity of function or clinical use. Each group was then used to create independent univariate or multivariable binary logistic regression models with DR as the end-point. A single Wald test p-value was derived for each model using *wald*.*test* from the *aod* package. ROC analysis was performed using *pROC*. McFadden’s and Nagelkerke’s pseudo-R^2^ were derived via the *pscl* and *rms* packages, respectively.

## RESULTS

### Study cohort

In NHANES 2007-8, retinal imaging data is available for 3863 individuals, demographics data is available for 10149 individuals, and laboratory data is available for between 394 and 9307 individuals, depending on the individual laboratory dataset in NHANES (see **Figure 1** footnote). After aligning all data and filtering for those who had diabetes by our classification, 1025 individuals remained in our dataset. The selection process is illustrated in **Figure 1**, while **Table 1** provides an overview of the demographics of these individuals.

**Table 1.**
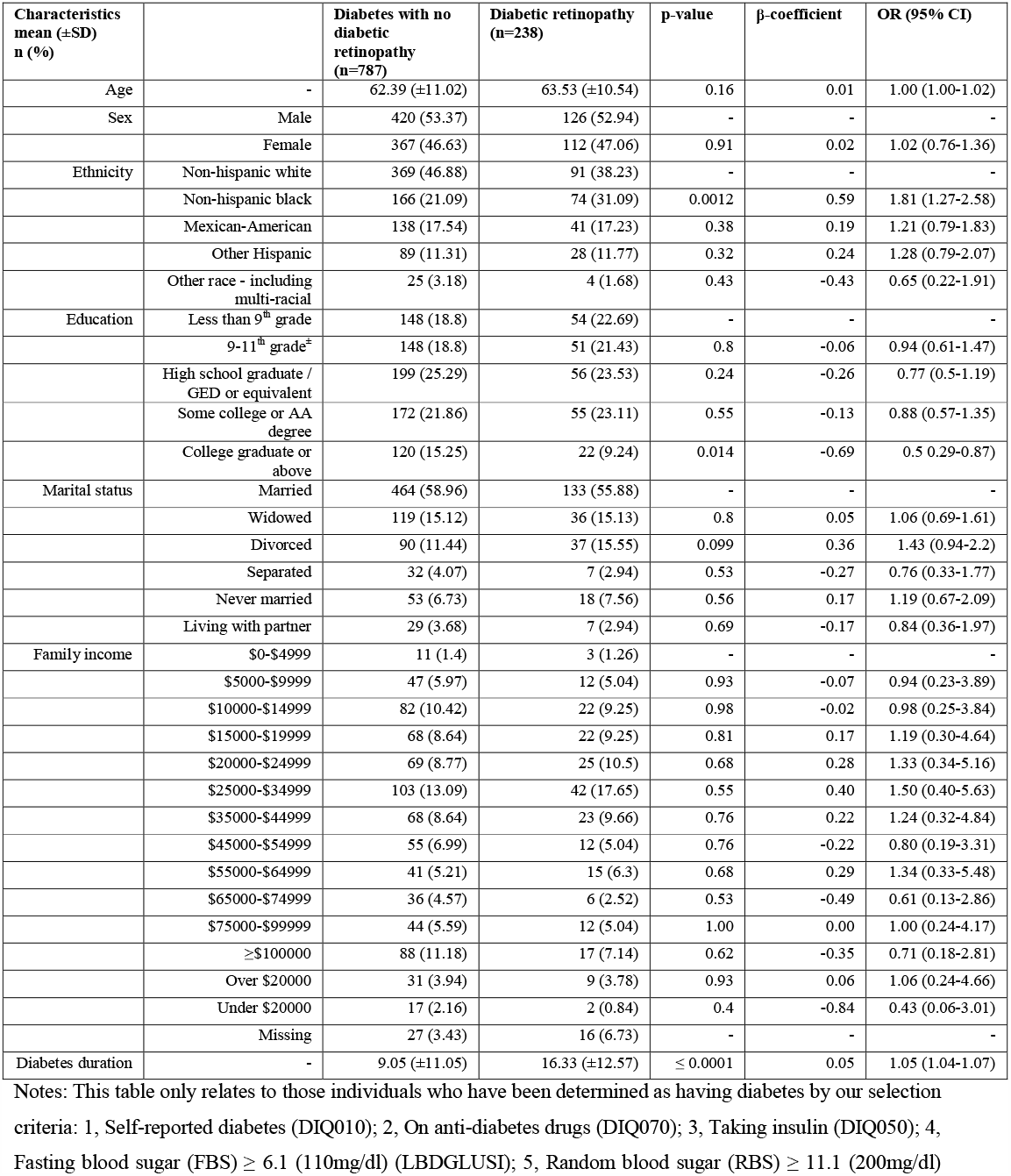
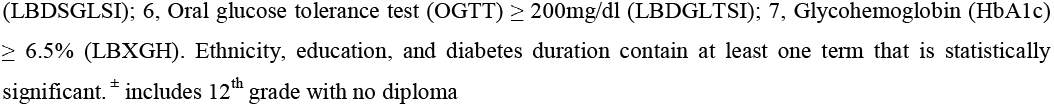
Demographic overview of study cohort.

| Characteristics<br>mean (±SD)<br>n (%) | | Diabetes with no<br>diabetic<br>retinopathy<br>(n=787) | Diabetic retinopathy<br>(n=238) | p-value | β-coefficient | OR (95% CI) |
| --- | --- | --- | --- | --- | --- | --- |
| Age | - | 62.39 (±11.02) | 63.53 (±10.54) | 0.16 | 0.01 | 1.00 (1.00-1.02) |
| Sex | Male | 420 (53.37) | 126 (52.94) | - | - | - |
|  | Female | 367 (46.63) | 112 (47.06) | 0.91 | 0.02 | 1.02 (0.76-1.36) |
| Ethnicity | Non-hispanic white | 369 (46.88) | 91 (38.23) | - | - | - |
|  | Non-hispanic black | 166 (21.09) | 74 (31.09) | 0.0012 | 0.59 | 1.81 (1.27-2.58) |
|  | Mexican-American | 138 (17.54) | 41 (17.23) | 0.38 | 0.19 | 1.21 (0.79-1.83) |
|  | Other Hispanic | 89 (11.31) | 28 (11.77) | 0.32 | 0.24 | 1.28 (0.79-2.07) |
|  | Other race - including<br>multi-racial | 25 (3.18) | 4 (1.68) | 0.43 | -0.43 | 0.65 (0.22-1.91) |
| Education | Less than 9<sup>th</sup> grade | 148 (18.8) | 54 (22.69) | - | - | - |
|  | 9-11<sup>th</sup> grade<sup>±</sup> | 148 (18.8) | 51 (21.43) | 0.8 | -0.06 | 0.94 (0.61-1.47) |
|  | High school graduate /<br>GED or equivalent | 199 (25.29) | 56 (23.53) | 0.24 | -0.26 | 0.77 (0.5-1.19) |
|  | Some college or AA<br>degree | 172 (21.86) | 55 (23.11) | 0.55 | -0.13 | 0.88 (0.57-1.35) |
|  | College graduate or<br>above | 120 (15.25) | 22 (9.24) | 0.014 | -0.69 | 0.5 (0.29-0.87) |
| Marital status | Married | 464 (58.96) | 133 (55.88) | - | - | - |
|  | Widowed | 119 (15.12) | 36 (15.13) | 0.8 | 0.05 | 1.06 (0.69-1.61) |
|  | Divorced | 90 (11.44) | 37 (15.55) | 0.099 | 0.36 | 1.43 (0.94-2.2) |
|  | Separated | 32 (4.07) | 7 (2.94) | 0.53 | -0.27 | 0.76 (0.33-1.77) |
|  | Never married | 53 (6.73) | 18 (7.56) | 0.56 | 0.17 | 1.19 (0.67-2.09) |
|  | Living with partner | 29 (3.68) | 7 (2.94) | 0.69 | -0.17 | 0.84 (0.36-1.97) |
| Family income | \$0-\$4999 | 11 (1.4) | 3 (1.26) | - | - | - |
| | \$5000-\$9999 | 47 (5.97) | 12 (5.04) | 0.93 | -0.07 | 0.94 (0.23-3.89) |
| | \$10000-\$14999 | 82 (10.42) | 22 (9.25) | 0.98 | -0.02 | 0.98 (0.25-3.84) |
| | \$15000-\$19999 | 68 (8.64) | 22 (9.25) | 0.81 | 0.17 | 1.19 (0.30-4.64) |
| | \$20000-\$24999 | 69 (8.77) | 25 (10.5) | 0.68 | 0.28 | 1.33 (0.34-5.16) |
| | \$25000-\$34999 | 103 (13.09) | 42 (17.65) | 0.55 | 0.40 | 1.50 (0.40-5.63) |
| | \$35000-\$44999 | 68 (8.64) | 23 (9.66) | 0.76 | 0.22 | 1.24 (0.32-4.84) |
| | \$45000-\$54999 | 55 (6.99) | 12 (5.04) | 0.76 | -0.22 | 0.80 (0.19-3.31) |
| | \$55000-\$64999 | 41 (5.21) | 15 (6.3) | 0.68 | 0.29 | 1.34 (0.33-5.48) |
| | \$65000-\$74999 | 36 (4.57) | 6 (2.52) | 0.53 | -0.49 | 0.61 (0.13-2.86) |
| | \$75000-\$99999 | 44 (5.59) | 12 (5.04) | 1.00 | 0.00 | 1.00 (0.24-4.17) |
| | ≥\$100000 | 88 (11.18) | 17 (7.14) | 0.62 | -0.35 | 0.71 (0.18-2.81) |
| | Over \$20000 | 31 (3.94) | 9 (3.78) | 0.93 | 0.06 | 1.06 (0.24-4.66) |
| | Under \$20000 | 17 (2.16) | 2 (0.84) | 0.4 | -0.84 | 0.43 (0.06-3.01) |
|  | Missing | 27 (3.43) | 16 (6.73) | - | - | - |
| Diabetes duration | - | 9.05 (±11.05) | 16.33 (±12.57) | ≤ 0.0001 | 0.05 | 1.05 (1.04-1.07) |

Notes: This table only relates to those individuals who have been determined as having diabetes by our selection criteria: 1, Self-reported diabetes (DIQ010); 2, On anti-diabetes drugs (DIQ070); 3, Taking insulin (DIQ050); 4, Fasting blood sugar (FBS) ≥ 6.1 (110mg/dl) (LBDGLUSI); 5, Random blood sugar (RBS) ≥ 11.1 (200mg/dl)
(LBDSGLSI); 6, Oral glucose tolerance test (OGTT) ≥ 200mg/dl (LBDGLTSI); 7, Glycohemoglobin (HbA1c) ≥ 6.5% (LBXGH). Ethnicity, education, and diabetes duration contain at least one term that is statistically significant. ± includes 12<sup>th</sup> grade with no diploma

For our replication cohort, NHANES 2005-6, we prepared laboratory data following the same filter criteria as NHANES 2007-8 and produced a final dataset of 2459 individuals, among which 637 (with retinopathy, 176; no retinopathy, 461) had diabetes.

### Retinal lesions of diabetic retinopathy

To help validate our methodology and cohort selection, we aimed to determine retinal lesions that define DR. To this end, we identified 9 retinal lesions in NHANES 2007-8 that were statistically significantly associated with DR and survived to p-value adjustment for false discovery (**Table 2**). The top lesions were retinal microaneurysms (p≤0.0001), followed by retinal hard exudates (typically due to lipoprotein deposition in the retina and may be associated with macular edema) (p≤0.0001). Other key lesions at p≤0.0001 were retinal soft exudate (now termed cotton wool spots), retinal blot hemorrhages, intraretinal microvascular abnormalities (IRMA), and macular edema. In NHANES, retinal microaneurysms and retinal blot hemorrhages are encoded to be mutually exclusive, i.e., an individual is recorded as having retinal microaneurysms only when not accompanied with retinal blot hemorrhages, and *vice-versa* (**Table 2**).

**Table 2.** Retinal lesions that constitute diabetic retinopathy outcome.

| Retinal co-morbidity | n (%) | β-coefficient | OR (95% CI) | p-value | FDR-adjusted p-value |
| --- | --- | --- | --- | --- | --- |
| Retinal microaneurysms only, worse eye | 129 (12.59) | 5.67 | 288.87 (127.66-653.68) | ≤ 0.0001 | ≤ 0.0001 |
| Retinal hard exudate, worse eye | 86 (8.39) | 3.72 | 41.34 (20.31-84.17) | ≤ 0.0001 | ≤ 0.0001 |
| Retinal blot hemorrhages, worse eye | 47 (4.59) | 3.36 | 28.71 (14.11-58.42) | ≤ 0.0001 | ≤ 0.0001 |
| Retinal soft exudate, worse eye | 76 (7.41) | 4.2 | 66.49 (26.44-167.21) | ≤ 0.0001 | ≤ 0.0001 |
| IRMA, worse eye | 62 (6.05) | 2.85 | 17.36 (9.06-33.26) | ≤ 0.0001 | ≤ 0.0001 |
| Macular edema, worse eye | 51 (4.98) | 3.88 | 48.24 (17.18-135.44) | ≤ 0.0001 | ≤ 0.0001 |
| Retinal fibrous proliferation, worse eye | 19 (1.85) | 3.41 | 30.19 (6.92-131.67) | ≤ 0.0001 | ≤ 0.0001 |
| Macular edema in center, worse eye | 26 (2.54) | 4.53 | 92.33 (12.45-684.9) | ≤ 0.0001 | ≤ 0.0001 |
| Retinal new vessels elsewhere, worse eye | 15 (1.46) | 3.12 | 22.68 (5.08-101.23) | ≤ 0.0001 | 0.0003 |

Notes: Lesions are taken from the NHANES *ophthalmology - retinal imaging* (OPXRET\_E) dataset. Only lesions with FDR-adjusted $p \leq 0.05$ are listed. Soft exudate is now termed cotton wool spots.

### Univariate logistic regression analysis

In total, 6 variables reached statistical significance in the unadjusted univariate analysis, 11 after adjustment for age, 2 after adjustment for ethnicity, and 7 for diabetes duration (**Supplementary Figure 1; Table 3**). Glycohemoglobin (HbA1c) was the only variable that was statistically significant in both the unadjusted and adjusted analyses. Other risk variables of note that reached statistical significance in the unadjusted analysis included serum glucose (mmol/L) (i.e., RBS), osmolality (mmol/Kg), urinary albumin (mg/L), and fasting glucose (mmol/L) (i.e., FBS). The only protective variable, i.e., negatively associated, was hemoglobin (g/dL). These variables indicate suboptimal diabetes control, abnormal kidney function and presence of anemia as risk factors for DR. There was evidence of co-correlation among these statistically significant variables from the unadjusted analysis (**Supplementary Figure 2**).

**Table 3.**
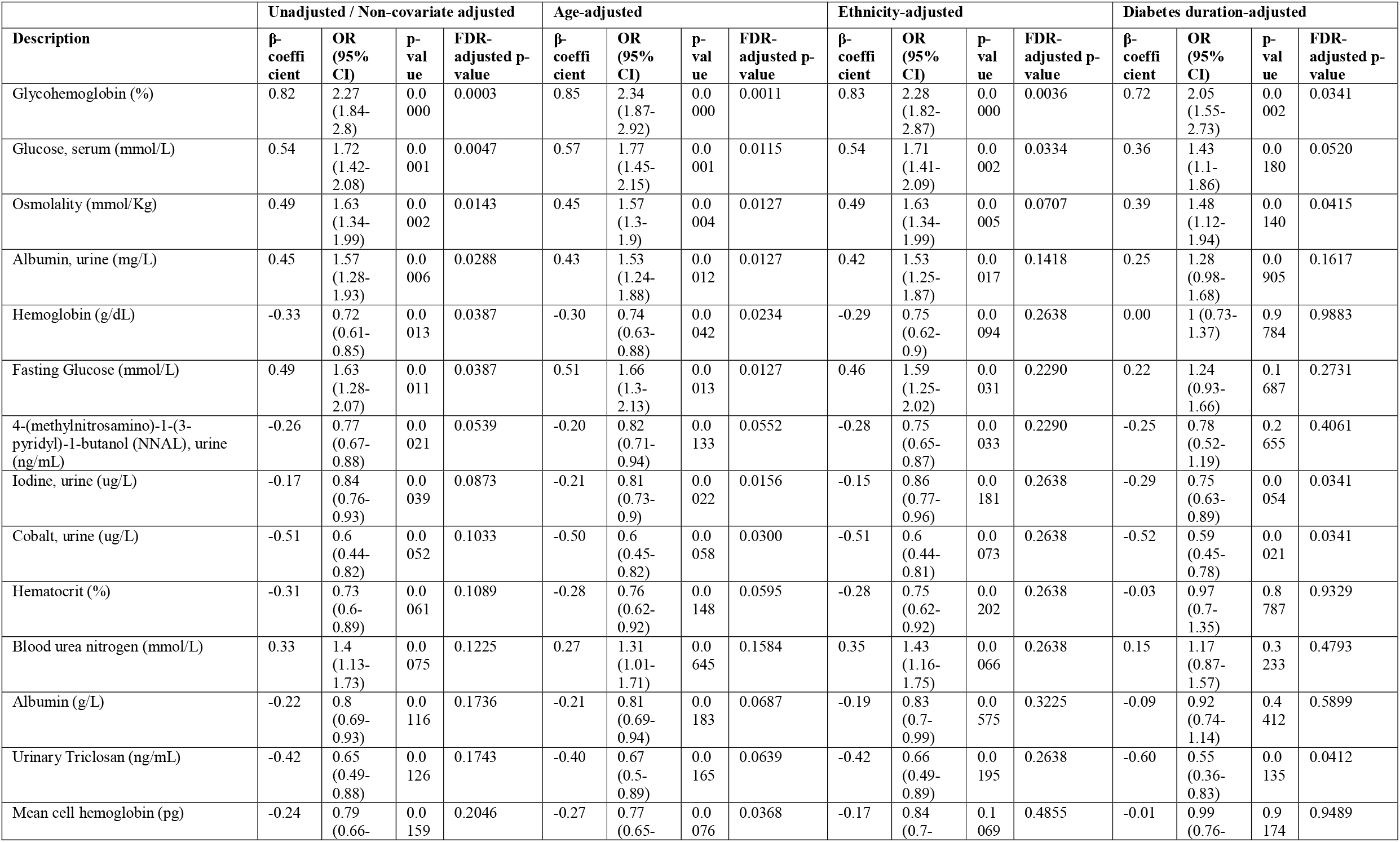
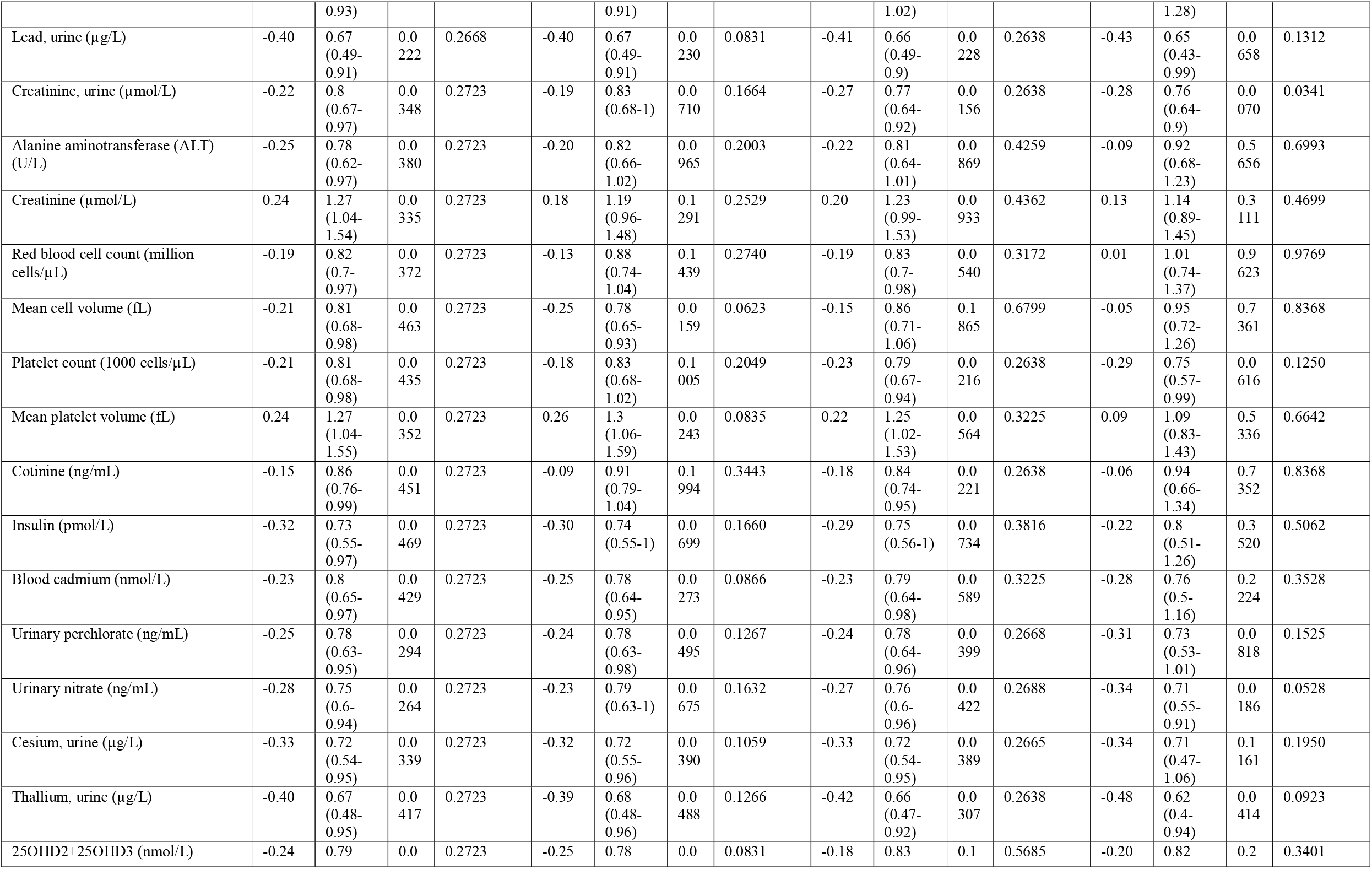
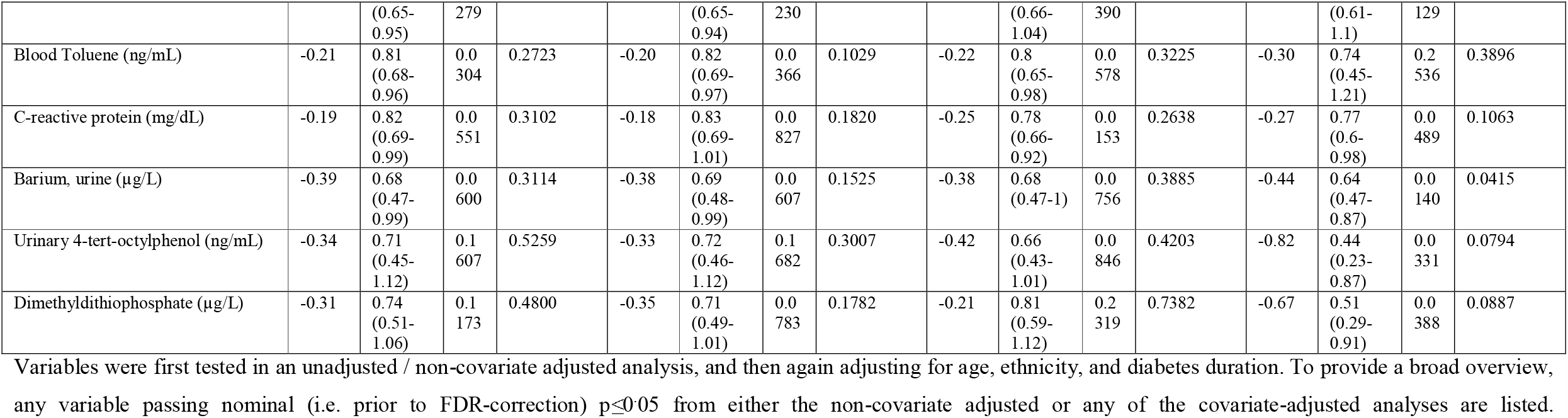
Laboratory variables associated with retinopathy in individuals with diabetes.

Interestingly, after adjustment for diabetes duration, the following variables reached statistical significance: HbA1c (%), osmolality (mmol/Kg), urinary iodine (µg/L), urinary cobalt (µg/L), urinary triclosan (ng/mL), urinary creatinine (µmol/L), and urinary barium (µg/L).

### Principal component analysis

Unsupervised PCA using all laboratory variables revealed that 59 PCs could account for 80% or more variation in the dataset. Eighteen PCs were statistically significantly associated with DR at p≤0.05 via independent binomial regression models testing each PC (**Supplementary Table 2**). The top variables responsible for variation along these PCs included measures of blood glucose (HbA1c, random blood glucose and fasting blood glucose), kidney function markers (urinary albumin, blood urea nitrogen [BUN]), hematological markers (hematocrit, hemoglobin, red blood cell distribution width), inflammatory markers (CRP), white blood cell count, urinary nitrates, segmented neutrophil count), and toxic elements (urinary beryllium and cotinine) among others – these PCs were also statistically significantly correlated to microaneurysms, the previously-identified top retinal lesion, and the covariates used during univariate testing **(Supplementary Figure 3**). Through ROC analysis, these 18 PCs achieved AUC 0.796 (95% CI: 0.761-0.832).

### Penalized regression model

We fitted an unbiased elastic-net penalized regression model to the laboratory variables and cross-validated it 100x. The model selected 9 variables whose coefficients were not shrunk to zero: urinary albumin, BUN, urinary cobalt, CRP, HbA1c, blood osmolality, serum potassium, systolic blood pressure, and urinary nitrate (**Figure 2**). Of note, these measurements mainly represent diabetes and blood pressure control and kidney function. This model had an accuracy of 78.51% and AUC 0.74 (95% CI: 0.72-0.77) when predicted on the same dataset on which the model was produced.

**Figure 2.**
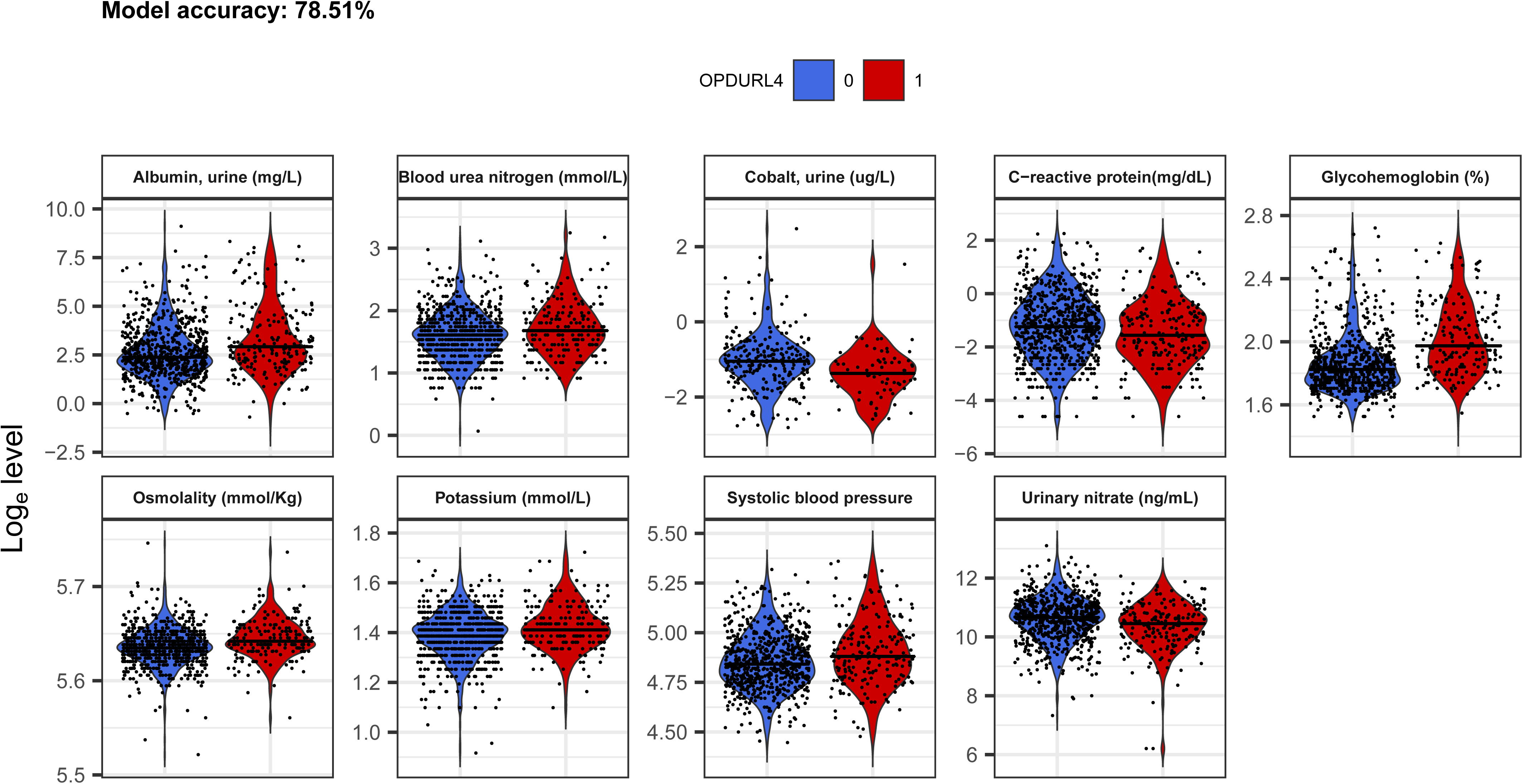
Penalized regression-selected variables. Variables were selected from a 100x cross-validated model with α=0.5. Final variable selection was based on coefficients not shrunk to 0. Model accuracy was determined to be 78.4% accuracy and AUC 0.71 (95% CI: 0.65-0.77).

### RandomForest™ classification model

From our RandomForest™ model, HbA1c was the single best predictor of DR (mean decrease accuracy, 31.94%; Gini, 21.75) (**Table 4**). However, other notable variables of appreciable accuracy were markers of diabetes control (FBS, RBS) inflammation (CRP), kidney function (potassium, BUN, creatinine and urinary albumin), hematological markers (hematocrit), systolic blood pressure, among others. The overall accuracy of the model on the validation cohort was 78.4% and AUC 0.71 (95% CI: 0.65-0.77).

**Table 4.** RandomForest™-selected variables (features).

| Marker | Group | Mean decrease accuracy | Mean decrease Gini |
| --- | --- | --- | --- |
| Glycohemoglobin (%) | Diabetes status | 31.93719324 | 21.75225602 |
| C-reactive protein (mg/dL) | Immune markers | 11.51047187 | 10.1975135 |
| Potassium (mmol/L) | Renal function | 11.44142126 | 8.198116194 |
| Albumin, urine (mg/L) | Renal function | 8.220187056 | 10.21346621 |
| Monocyte number (1000 cells/uL) | Immune markers | 7.663589246 | 3.957711466 |
| Osmolality (mmol/Kg) | Renal function | 7.510989556 | 5.33445026 |
| White blood cell count (1000 cells/uL) | Immune markers | 7.440157644 | 4.405168072 |
| Blood urea nitrogen (mmol/L) | Renal function | 7.224789174 | 5.073947519 |
| Segmented neutrophils num (1000 cell/uL) | Immune markers | 7.020397225 | 4.067402271 |
| Fasting Glucose (mmol/L) | Diabetes status | 6.563694988 | 2.826827797 |
| Red cell distribution width (%) | Hematocrit | 6.138538515 | 4.947185792 |
| Urinary nitrate (ng/mL) | Renal function | 5.899174386 | 5.844258103 |
| Glucose, serum (mmol/L) | Diabetes status | 5.85339684 | 4.77357191 |
| 2-hydroxyphenanthrene (ng/L) | Toxins / Metals | 4.028082549 | 2.307374348 |
| MCHC (g/dL) | Hematocrit | 3.936015913 | 3.896804592 |
| Creatinine (μmol/L) | Renal function | 3.530916132 | 3.747500758 |
| Mono-2-ethyl-5-carboxypentyl phthalate | Toxins / Metals | 2.996581682 | 2.212151134 |
| Blood Nitromethane (pg/mL) | Toxins / Metals | 2.936160917 | 3.31634662 |
| Phosphorus (mmol/L) | Toxins / Metals | 2.819084205 | 3.768671211 |
| Total Cholesterol (mmol/L) | Sterols | 2.448578413 | 3.833301666 |
| Enterodiol (ng/mL) | Sterols | 2.401651721 | 2.722762228 |
| Hematocrit (%) | Hematocrit | 2.364741874 | 4.841281382 |
| Mono-n-octyl phthalate (ng/mL) | Toxins / Metals | 2.215508752 | 0.231647322 |
| Mean cell hemoglobin (pg) | Hematocrit | 2.183482824 | 4.292282119 |
| Gamma glutamyl transferase (U/L) | Liver Function | 1.989695745 | 4.015070221 |
| Systolic blood pressure | Blood pressure | 1.892815461 | 8.85430752 |
| Blood o-Xylene (ng/mL) | Toxins / Metals | 1.670964308 | 3.708248225 |
| Lactate dehydrogenase LDH (U/L) | Liver Function | 1.593869272 | 4.231659273 |
| 9-hydroxyfluorene (ng/L) | Toxins / Metals | 1.430814333 | 2.206778568 |
| Cholesterol (mmol/L) | Sterols | 1.201366974 | 4.025207689 |
| 3-hydroxyphenanthrene (ng/L) | Toxins / Metals | 1.00569817 | 1.63630888 |

Notes: The model was initially trained on all laboratory variables in an unsupervised fashion, with Kappa-based model tuning to select the optimum values for ‘mtry’ (the ideal number of variables to randomly sample) and ‘ntrees’ (the ideal number of trees). Only variables contributing >1% mean decrease in accuracy from the initial model were retained, followed by recursive steps to remove low-informative variables. Variables are manually assigned to groups based on similar organ function or other characteristic. The Gini importance measure relates to the ‘splitting’ criterion that is employed in classification trees, and it is known to be less biased for continuous variables (26), which naturally have more splitting points compared to categorical variables. ‘Group’ is manually curated.

### Replication cohort

In the NHANES 2005-6 replication cohort, we performed penalized regression and RandomForest™ in the same way as per the 2007-8 cohort. Our penalized regression model identified urinary albumin (mg/L), cockroach IgE antibody (kU/L), HbA1c (%), hemoglobin (g/dL), and urinary nitrate (ng/mL), with a model accuracy of 73.16% and AUC 0.76 (95% CI: 0.73-0.78). RandomForest™ identified HbA1c (%) as the variable contributing most to accuracy (mean decrease 16.96%), with many other variables contributing appreciable accuracy to the overall model (**Supplementary Table 3**) - the overall model accuracy was 72.98% and AUC 0.68 (95% CI: 0.61-0.75).

### Final clinical risk models

The 31 features identified by RandomForest™ (**Table 4**) were grouped into different categories of blood tests according to diabetes status, hematocrit values, blood pressure (BP), immune markers, renal function, sterols, toxins and metals, and liver function. When modeled against DR outcome, each group varied in performance; diabetes tests alone achieved AUC 0.72 (95% CI: 0.68-0.76). A final clinical risk model comprising diabetes tests, BP, renal and liver function tests, hematocrit values, circulating sterols and immune markers achieved AUC 0.84 (95% CI: 0.78-0.9) (p=0.00013) (Nagelkerke R^2^ 0.36) (**Table 5**; **Figure 3**).

**Figure 3.**
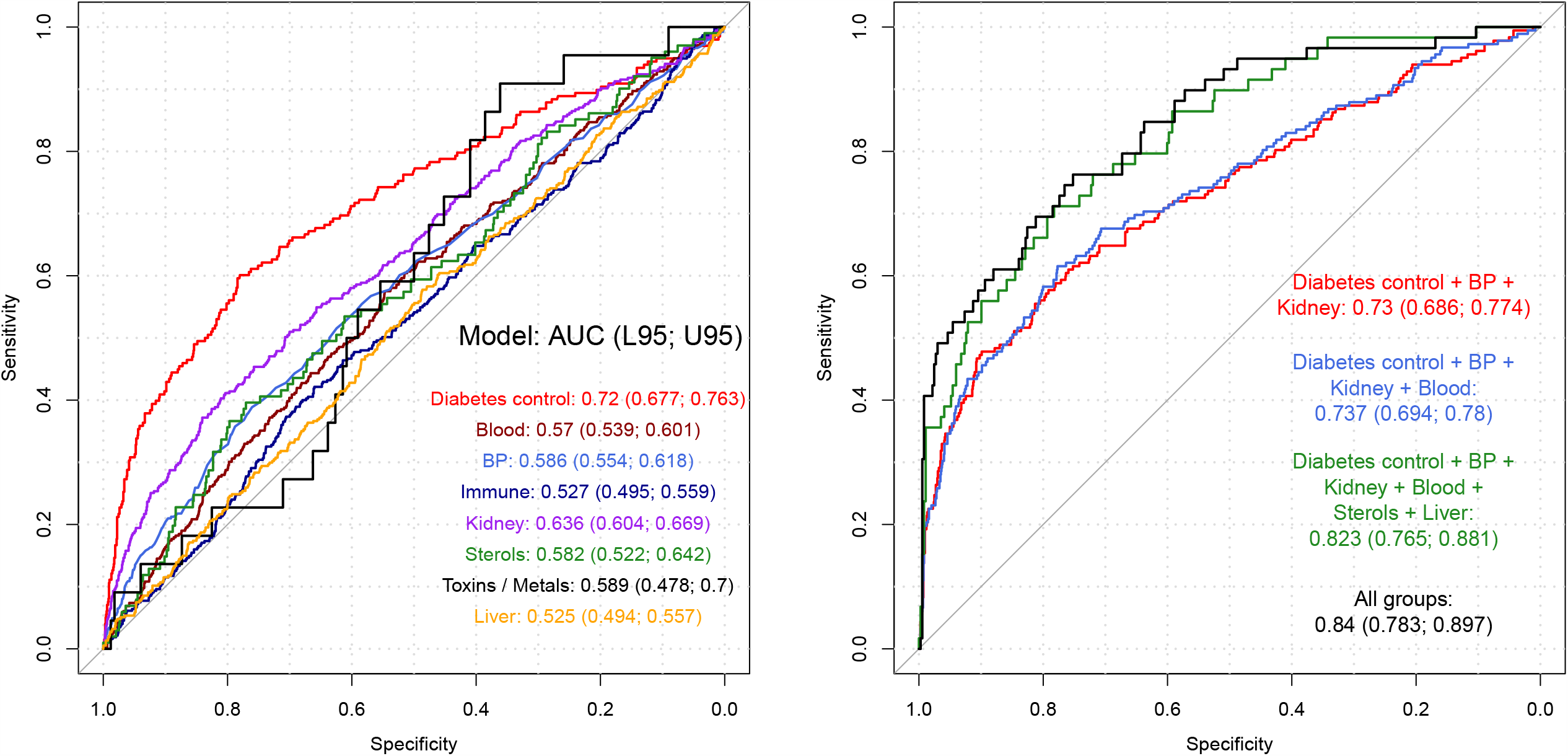
Final clinical risk models. Features from RandomForest™ were grouped logically based on similar function or clinical use (**Table 4**) and then tested independently in a univariate or multivariate regression model against DR outcome. A final risk model including markers of hypertension, hypercholesterolemia, renal and liver function tests, and hematocrit achieved AUC 0.84 (0.78-0.9).

**Table 5.** Final clinical risk models.

| Model | Wald test p-value | McFadden R<sup>2</sup> | Nagelkerke R<sup>2</sup> | AUC (95% CI) |
| --- | --- | --- | --- | --- |
| Diabetes Status | ≤ 0.0001 | 0.102 | 0.142 | 0.72 (0.677-0.763) |
| Hematocrit | 0.0023 | 0.007 | 0.009 | 0.57 (0.539-0.601) |
| Blood Pressure (BP) | ≤ 0.0001 | 0.017 | 0.024 | 0.586 (0.554-0.618) |
| Immune Markers | 0.42 | 0.002 | 0.002 | 0.527 (0.495-0.559) |
| Renal function tests (renal) | ≤ 0.0001 | 0.039 | 0.054 | 0.636 (0.604-0.669) |
| Sterols (include cholesterol) | 0.055 | 0.012 | 0.017 | 0.582 (0.522-0.642) |
| Toxins / Metals | 0.94 | 0.029 | 0.041 | 0.589 (0.478-0.7) |
| Liver function tests | 0.07 | 0.002 | 0.003 | 0.525 (0.494-0.557) |
| Diabetes control + BP + Renal function | $\leq 0.0001$ | 0.13 | 0.18 | 0.73 (0.686-0.774) |
| Diabetes control + BP + renal function + Hematocrit | $\leq 0.0001$ | 0.135 | 0.184 | 0.737 (0.694-0.78) |
| Diabetes control + BP + renal function + Hematocrit + Sterols + Liver function | $\leq 0.0001$ | 0.238 | 0.315 | 0.823 (0.765-0.881) |
| All groups ± | 0.00013 | 0.272 | 0.355 | 0.84 (0.783-0.897) |

± The only toxins / metal included was Phosphorus (mmol/L) – others filtered out due to high missingness (>50%), resulting in difficulty fitting model.

## DISCUSSION

This EWAS of NHANES 2007-8 data with DR outcomes in individuals with diabetes included an unbiased feature selection approach based on a rudimentary univariate regression enabled for compute parallelization, PCA, penalized regression, and RandomForest™ of a large number of laboratory parameters. In contrast, epidemiological studies are typically conducted based on pre-conceived hypotheses and involve a single or just a few variables. These methods can be scaled to datasets of any size and therefore provide ways of working with large clinical and epidemiological datasets for the purpose of searching for novel hypotheses that could then lead to further focused investigations.

In our rudimentary approach, which is ultimately running many univariate models in a parallelized fashion, HbA1c was the only variable to reach statistical significance after adjusted for age, ethnicity, and diabetes duration. The relationship between HbA1c and DR has been explored extensively and was selected as the strongest risk factor in every approach we undertook, with a mean decrease accuracy of 31.94% via RandomForest™.

Our penalized regression and RandomForest™ algorithms also identified an association between elevated systolic blood pressure —but not diastolic— and DR (mean decrease accuracy, 1.9%), again confirming literature (10, 28-30). Further risk variables identified by both penalized regression and RandomForest™ were renal function tests including BUN, urinary albumin, potassium, osmolality, and urinary nitrate. These confirm the strong association of DR with markers of impaired kidney function. Other known risk factors that contributed higher up in the ranking order include hematocrit (%) and cholesterol. Although HbA1c has the strongest association with DR, our study highlight how the addition of other clinical parameters, e.g., from renal and liver function and hematocrit can increase the sensitivity and specificity of predicting DR outcome, with our final clinical risk model achieving AUC 0.83 (95% CI: 0.77-0.89) (p=0.00012) (Nagelkerke R^2^ 0.33), higher than any traditional diabetes control parameter in isolation or in combination.

The EWAS methodology and our RandomForest™ approach of non-targeted recursive feature also indicates a small contribution from toxins and metals, including 3-hydroxyphenanthrene, 9-hydroxyfluorene, phthalates, blood o-Xylene, and blood nitromethane. Therefore, retina may be a target tissue for environmental contamination. Some of the associations provide directions to future mechanistic research in DR. For example, we found that retinal microaneurysms (FDR-adjusted p≤0.0001), the most statistically significant retinal lesions in individuals with DR, is already correlated with some of the variables such as HbA1c, CRP, BUN, beryllium, and hematocrit, suggesting early effects. In contrast, increased urinary cobalt, triclosan and barium became significant only when adjusted for duration of diabetes. Most of these parameters are also linked to risk of allergies and lung disease, an association that has not been previously explored systematically.

As this is a cross-sectional study, a cause-effect relation cannot be established. Moreover, we are unable to rule out any confounding effects of any unmeasured factors. On the other hand, the main strength of the study is the use of the well characterised NHANES cohort in whom standardised protocols were used to measure laboratory parameters. We are not aware of any other association studies in DR where over 400 laboratory parameters were analysed simultaneously to develop multiple models. As the top variables of all four data driven agnostic models were similar, we also believe our findings are generalisable.

## CONCLUSION

We confirm that DR is a complex disease and that the already established risk factors contribute significantly to the risk models of DR, with HbA1c being the strongest risk factor. Although our model provides an accuracy of approximately 80%, it also provides mechanistic insights into future research on DR including interrogating the interaction of low-ranking risk factors with more established factors in the models and highlights need to explore epigenetic screens to gauge better how risk factors influence gene expression. Most importantly, the study reinforces the need to control known risk factors of DR, especially hyperglycemia.

## Data Availability

Data is available from The National Health and Nutrition Examination Survey (NHANES - https://wwwn.cdc.gov/Nchs/Nhanes/)

## ACKNOWLEDGEMENTS

We thank the many thousands of NHANES study participants who, over the course of decades, have provided valuable information for epidemiological studies.

## AUTHOR CONTRIBUTIONS

KB, SG and SS conceived and designed the study. KB, SG and YL analysed the data. KB is the study data guarantor. All authors interpreted the results and reviewed the manuscript. KB had full access to all of the data and takes responsibility for the integrity of the data and the accuracy of data analysis. All authors read and approved the final manuscript.

